# ECG abnormalities are strongly associated with incident heart failure events in low-risk individuals using the PREVENT HF risk equations

**DOI:** 10.64898/2026.05.18.26353538

**Authors:** Mouhammad J Alawad, Elsayed Z. Soliman, Todd M. Brown, Oluwasegun P. Akinyelure, Yulia Khodneva, Hugo G. Quezada-Pinedo, Mohamed A. Mostafa, Mohan Satish, Parag Goyal, Orysya Soroka, Monika M. Safford

## Abstract

**Background:** Resting electrocardiogram (ECG) are associated with heart failure (HF) events, even though it is not currently recommended in risk assessment. The role of ECG abnormalities is not clear in identifying at risk individuals.

**Objective:** To examine the association between ECG abnormalities and incident HF events according to the 2023 Predicting Risk of Cardiovascular Disease Events (PREVENT) HF equation. And identify a subgroup of individuals who are misclassified as being at low risk and may benefit from primary intervention measures.

**Design:** Secondary data analysis from the REasons for Geographic And Racial Differences in Stroke (REGARDS) prospective cohort, including study participants without baseline HF.

**Exposure:** ECG abnormalities were classified by Minnesota Code (MC) as normal, any minor, or any major abnormality at baseline (2003-2007).

**Outcome:** Participants were followed for expert adjudicated incident HF hospitalizations/ deaths through December 31, 2021.

**Results:** Among 20,923 participants (mean age at baseline of 63.6 years, 53.7% were female), 26.0% of the sample was classified as low risk (<3%), 17.5% as borderline risk (3-<5%), 27.5% as intermediate risk (5%-<10%), and 29.0% as high risk (≥10%). Overall, 43.8% had normal ECG, 41.7% had at least one minor abnormality, and 14.5% had at least one major abnormality. HF events occurred in 3.3% of the sample with normal ECG, 6.2% of those with any minor abnormality, and 13.2% of those with any major ECG abnormality. Compared to those without ECG abnormality, the adjusted HR for incident HF was 1.56 (95% CI 1.35-1.80) for any minor abnormality and 2.56 (2.18-3.00) for any major abnormality. 43.5% of the population were in the less than 5% risk by PREVENT among whom 45.8% had any ECG abnormalities. The fully adjusted HR for only minor ECG abnormalities in the <3% was 1.47 (95% CI 0.72-3.01), and the fully adjusted HR for any major ECG abnormality was 5.22 (95% CI 2.42-11.30). In the borderline risk group, the fully adjusted HR for only minor ECG abnormalities was 1.37 (95% CI 0.89 - 2.11), and the fully adjusted HR for any major ECG abnormality was stronger than the HR in the intermediate and high-risk groups; 3.05 (95% CI 1.85 - 5.03).

**Conclusion:** ECG abnormalities were common and associated with HF events across all PREVENT risk groups, especially in the low/borderline risk groups with major ECG abnormalities. ECG may be useful to identify at-risk individuals who would otherwise be misclassified as lower risk patients.

## INTRODUCTION

Heart failure (HF) is a growing public health problem; approximately 1 in 4 persons in the US will develop HF in their lifetime.^1^ By 2050, the prevalence of HF is projected to increase from 2.7% to 3.8% in all age groups and ethnic categories.^2^ HF mortality rates have been increasing since 2012,^1^ and 33.1% of cardiovascular disease (CVD) deaths are HF related.^3^ As the US population ages, so will the prevalence of HF and its concomitant health care related expenditures.^1,3^

Despite the advancement of medical and mechanical management of HF and heart transplantation, a preventive approach is needed to identify individuals at risk who could benefit from aggressive primary prevention. The 2022 American College of Cardiology (ACC)/American Heart Association (AHA)/Heart Failure Society of America guidelines recommend to use multivariable risk scores to estimate risk of HF, to use natriuretic peptide to screen for individuals at risk, and to refer at-risk individuals to specialized HF care for evidence-based strategies of HF prevention.^4^ No primary prevention trials have been conducted with a risk-based approach for HF; however, the Predicting Risk of Cardiovascular Disease Events (PREVENT) equations include risk stratification specifically for HF, and thus open the door for a new approach of identifying at risk individuals for targeted intervention.^5^

HF prevention strategies are most important for those at highest risk. This supports effort to develop and/or optimize available models that can correctly classify an individual’s HF risk profile. ECG is a widely available diagnostic tool, but few studies have examined an association between ECG abnormalities and incident HF, ^6–11^ and such studies are outdated, not based on systematic ECG classification (such as the Minnesota code or the Novacode), have small samples, and use older risk stratification scores. The role of ECG abnormalities is not clear in identifying at risk individuals. In this study, we sought to examine whether ECG abnormalities would identify a subgroup of individuals using the validated PREVENT equation, especially those who are considered low risk or borderline risk.

For this study, we used data from the large national bi-racial REasons for Geographic And Racial Differences in Stroke (REGARDS) cohort to examine the relationship between baseline ECG abnormalities and future incident HF hospitalization and HF mortality using the PREVENT HF equation for risk stratification. We hypothesized that ECG abnormalities are associated with risk of HF events in all HF risk groups as assessed by the 2023 PREVENT HF equations.

## METHODS

### Study Population

The REGARDS study is a prospective national biracial cohort designed to define regional and racial differences in stroke mortality. Details are provided elsewhere;^12^ in brief, from 2003 to 2007, 30,239 community-dwelling individuals aged 45 years and older were enrolled using commercially available lists. The sampling design over-recruited from the Southeastern Stroke Belt states, with 50% residing in Louisiana, Mississippi, Alabama, Georgia, North and South Carolina, Tennessee, and Arkansas, and 50% residing in the other contiguous US states. Within the Stroke Belt, one third of the sample resided in the Stroke Buckle, consisting of the coastal areas of North and South Carolina and Georgia. Self-reported race other than Black or white, inability to speak English, active cancer treatment, and inability to complete a survey and in-home exam were exclusions. Baseline data included a medical history and in-home exam that collected anthropometrics, blood and urine samples, and an ECG. Participants are contacted by telephone at 6-month intervals to detect potential CVD events with subsequent retrieval of medical records for adjudication by trained and calibrated clinician experts. Follow-up is ongoing. Written informed consent was obtained from all subjects, and the institutional review boards at all participating institutions approved the protocol.

For this study, participants free of HF were included using an algorithm to define an HF-free phenotype with negative predictive value of 99%.^13^

### Main exposure: ECG abnormalities

Electrocardiograms obtained during the in-home baseline visit were transmitted for centralized interpretation at the Epidemiological Cardiology Research Center (Wake Forest School of Medicine, Winston-Salem, NC, USA). All tracings were read by trained analysts blinded to clinical data and study outcomes. ECG findings were classified according to the Minnesota Code (MC).^14^ Participants with only minor abnormalities were categorized as having a minor ECG abnormality, whereas those with any major abnormality—regardless of the presence of coexisting minor abnormalities—were categorized as having a major ECG abnormality. Major ECG abnormalities included: Major ventricular conduction defect; definite myocardial infarction (defined as the presence of major Q wave abnormalities); possible myocardial infarction (defined as the presence of minor Q-QS wave plus major ST-T abnormalities); major isolated ST-T abnormalities; left ventricular hypertrophy plus major ST-T abnormalities; major atrioventricular conduction abnormalities; major QT prolongation (QTI≥116% or JTI if QRS ≥120 ms), pacemaker, and other major arrhythmias. Minor ECG abnormalities included: Minor isolated Q-QS waves; minor isolated ST-T abnormalities; high R waves; ST segment elevation; incomplete right bundle branch block; minor QT prolongation (QTI≥112% or JTI if QRS≥120 ms); short PR interval; left axis deviation; right axis deviation; frequent ventricular premature beats; and other minor abnormalities. For a detailed description of major and minor ECG abnormalities, see supplementary Table 2.

### Outcomes

The primary outcome was incident HF event, defined as a first-time HF hospitalization or HF death. HF was adjudicated based on a hospitalization with a clinical presentation consistent with HF, treatment consistent with HF, B-type natriuretic protein elevation, and imaging findings.^15^ HF mortality was defined as HF being the main underlying cause of death, adjudicated using all available information, including medical history, recent hospitalization, death certificates, interviews with next-of-kin, and the National Death Index.

### Covariates

Covariates were selected based on confounding and to account for study sampling (stroke belt, stroke buckle, neither). Covariates included sociodemographics (age, sex, annual household income, education), health behaviors (cigarette smoking, physical activity, adherence to a Mediterranean diet), chronic medical conditions and treatment (hypertension [HTN], treatment for HTN, statin treatment, diabetes, insulin treatment, chronic kidney disease as reflected in high albumin-to-creatinine ratio and estimated glomerular filtration rate [eGFR] using the 2021 CKD Epi equation, depressive symptoms), inflammation (high sensitivity C-reactive protein [CRP]), lipid biomarkers (total cholesterol, HDL cholesterol), anthropometrics (systolic blood pressure [BP], body mass index [BMI]), and family history of premature CVD.

## Statistical Analysis

We categorized the sample into the 3 ECG groups defined above (normal, only minor, and any major abnormalities). We used the extended PREVENT HF equation including the social deprivation index (SDI) to classify participants as low risk (<3%), borderline risk (3% - <5%), intermediate risk (5%-<10%), and high risk (≥10%) of experiencing a HF event in the next 10 years. The PREVENT equation included age, sex, cigarette smoking, total cholesterol, HDL cholesterol, systolic BP, HTN medication treatment, diabetes, eGFR, statins.^16^ Participants without valid information to calculate PREVENT risk were excluded from the study. We then compared the characteristics of the 3 ECG groups on sociodemographic factors, HF risk factors, and HF risk groups. We reported the proportion of each HF risk group that experienced any HF event.

Next, we constructed Kaplan-Meier (KM) survival curves for HF events classifying individuals into the 3 ECG groups. We used the log-rank test to determine the statistical significance of differences in the curves. We created KM curves for the whole sample as well as for each HF risk group separately.

We then examined the association between baseline ECG category and future HF events. We used an analogous approach for each analysis. We used Cox proportional hazards regression to examine the risk of HF events associated with ECG category. Participants censored at the date of an outcome, date of withdrawal from the study, date of death, or December 31, 2021, whichever came first. We first constructed Cox proportional hazards models to examine the age- and sex-adjusted model for the association between a HF event and minor or major ECG abnormality groups compared to the no ECG abnormality group, generating hazard ratios (HR) and 95% confidence intervals (CI). We then added sociodemographic, health behaviors, chronic medical conditions, inflammatory markers, and medications to the model and observed changes in the magnitude and directionality of the parameter estimate for the main exposure variable (ECG risk group variable). We did not adjust for covariates already embedded in the HF PREVENT base equation to avoid adjusting twice for the same components. We accounted for the competing risk of non-HF death using the method described by Fine and Gray.^17^ For all analyses described above, the proportional hazards assumption was examined using Schoenfeld residuals.

To examine differences by sex, age (<65 and ≥65 years), and HF risk classification group, we tested interaction terms and stratified models if the p-value for the interaction was <0.10. we performed stratified analysis according to HF risk groups.

We implemented multiple imputation of covariates by chained equations to minimize bias and maximize sample size. Variables with the largest amount of missing data included Mediterranean diet score (26.9%), family history of premature CVD (11.8%), and income (11.6%). All other covariates had less than 3% missing data. All tests were 2-tailed and p < 0.05 was used as the threshold for statistical significance except in the test for interactions, where the p-value was set at <0.10. Analyses were conducted using SAS version 9.4 (SAS Institute, Cary, NC) and Stata, version 19.5 (StataCorp, College Station, TX).

## RESULTS

### Sample characteristics

Out of the 30,239 members of the REGARDS cohort, 26,651 participants had no evidence of HF at baseline and had ECG information. The final cohort included 20,923 participants who had all the information for the HF PREVENT Base + SDI equation. Median follow-up time was 13.3 years (IQR 7.0-16.1). The mean age at baseline was 63.6 years and 53.7% were female, 38.4% self-identified as having Black race, 10.6% reported less than a high School education, and 43.7% reported annual household income <$35,000 (Table 1 and Supplementary Table 1). The prevalence of HTN was 71.3%, 55.3% were receiving anti-HTN medications, 20.3% had diabetes, and 28.6% were on statins. In the study sample, 9,163 (43.8%) individuals had a normal ECG, 8,722 (41.7%) had any minor abnormality, and 3,038 (14.5%) had any major ECG abnormality.

**Table 1:**
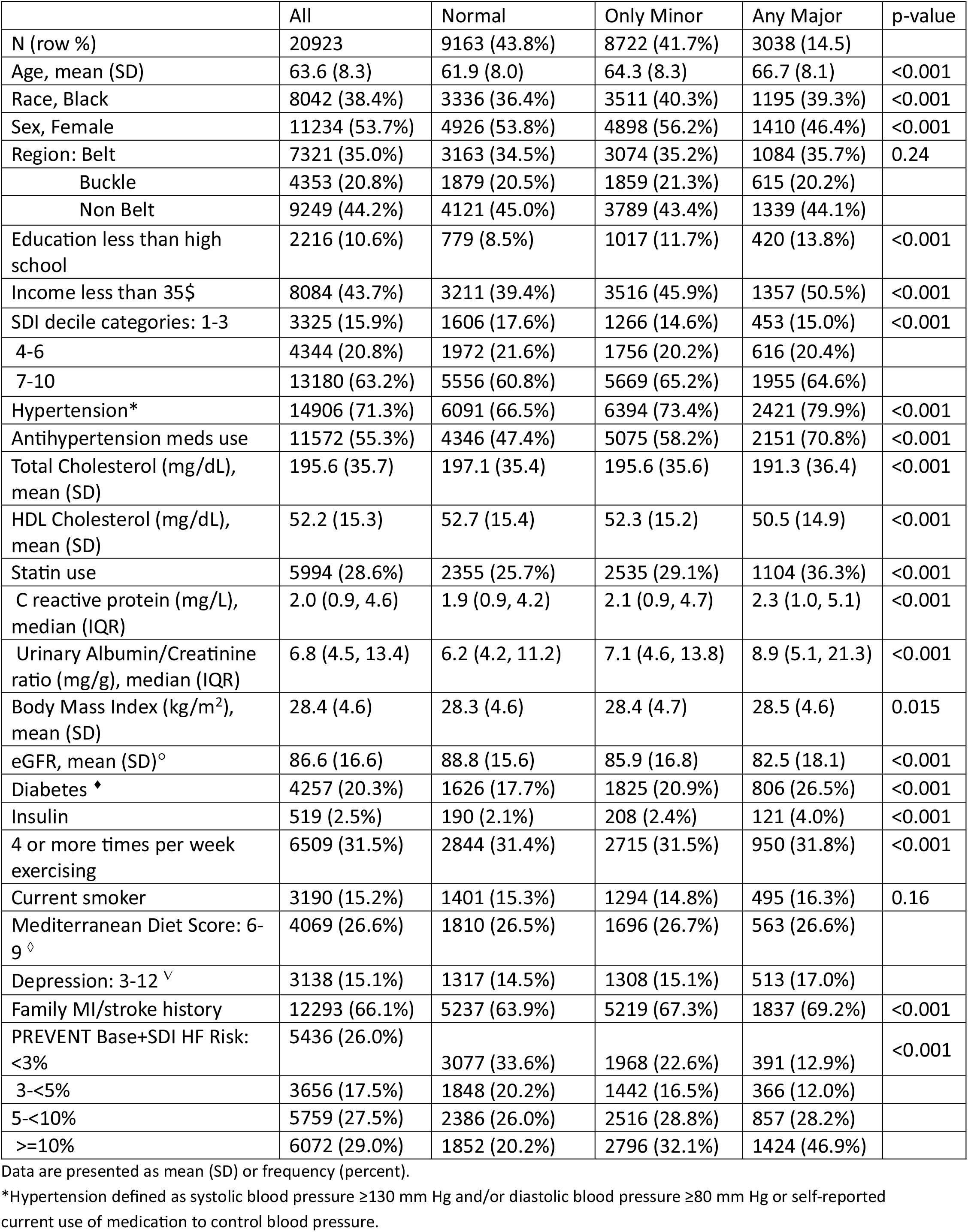
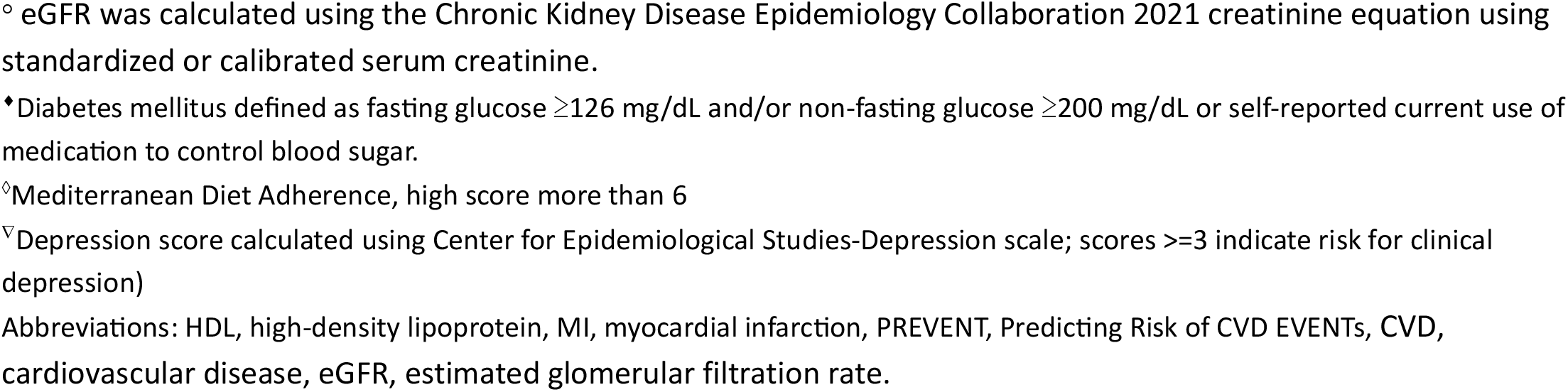
Baseline Characteristics of REGARDS Study Participants stratified by ECG categories.

|  | All | Normal | Only Minor | Any Major | p-value |
| --- | --- | --- | --- | --- | --- |
| N (row %) | 20923 | 9163 (43.8%) | 8722 (41.7%) | 3038 (14.5%) |  |
| Age, mean (SD) | 63.6 (8.3) | 61.9 (8.0) | 64.3 (8.3) | 66.7 (8.1) | <0.001 |
| Race, Black | 8042 (38.4%) | 3336 (36.4%) | 3511 (40.3%) | 1195 (39.3%) | <0.001 |
| Sex, Female | 11234 (53.7%) | 4926 (53.8%) | 4898 (56.2%) | 1410 (46.4%) | <0.001 |
| Region: Belt | 7321 (35.0%) | 3163 (34.5%) | 3074 (35.2%) | 1084 (35.7%) | 0.24 |
| Buckle | 4353 (20.8%) | 1879 (20.5%) | 1859 (21.3%) | 615 (20.2%) |  |
| Non Belt | 9249 (44.2%) | 4121 (45.0%) | 3789 (43.4%) | 1339 (44.1%) |  |
| Education less than high school | 2216 (10.6%) | 779 (8.5%) | 1017 (11.7%) | 420 (13.8%) | <0.001 |
| Income less than 35\$ | 8084 (43.7%) | 3211 (39.4%) | 3516 (45.9%) | 1357 (50.5%) | <0.001 |
| SDI decile categories: 1-3 | 3325 (15.9%) | 1606 (17.6%) | 1266 (14.6%) | 453 (15.0%) | <0.001 |
| 4-6 | 4344 (20.8%) | 1972 (21.6%) | 1756 (20.2%) | 616 (20.4%) |  |
| 7-10 | 13180 (63.2%) | 5556 (60.8%) | 5669 (65.2%) | 1955 (64.6%) |  |
| Hypertension* | 14906 (71.3%) | 6091 (66.5%) | 6394 (73.4%) | 2421 (79.9%) | <0.001 |
| Antihypertension meds use | 11572 (55.3%) | 4346 (47.4%) | 5075 (58.2%) | 2151 (70.8%) | <0.001 |
| Total Cholesterol (mg/dL), mean (SD) | 195.6 (35.7) | 197.1 (35.4) | 195.6 (35.6) | 191.3 (36.4) | <0.001 |
| HDL Cholesterol (mg/dL), mean (SD) | 52.2 (15.3) | 52.7 (15.4) | 52.3 (15.2) | 50.5 (14.9) | <0.001 |
| Statin use | 5994 (28.6%) | 2355 (25.7%) | 2535 (29.1%) | 1104 (36.3%) | <0.001 |
| C reactive protein (mg/L), median (IQR) | 2.0 (0.9, 4.6) | 1.9 (0.9, 4.2) | 2.1 (0.9, 4.7) | 2.3 (1.0, 5.1) | <0.001 |
| Urinary Albumin/Creatinine ratio (mg/g), median (IQR) | 6.8 (4.5, 13.4) | 6.2 (4.2, 11.2) | 7.1 (4.6, 13.8) | 8.9 (5.1, 21.3) | <0.001 |
| Body Mass Index (kg/m<sup>2</sup>), mean (SD) | 28.4 (4.6) | 28.3 (4.6) | 28.4 (4.7) | 28.5 (4.6) | 0.015 |
| eGFR, mean (SD)<sup>o</sup> | 86.6 (16.6) | 88.8 (15.6) | 85.9 (16.8) | 82.5 (18.1) | <0.001 |
| Diabetes * | 4257 (20.3%) | 1626 (17.7%) | 1825 (20.9%) | 806 (26.5%) | <0.001 |
| Insulin | 519 (2.5%) | 190 (2.1%) | 208 (2.4%) | 121 (4.0%) | <0.001 |
| 4 or more times per week exercising | 6509 (31.5%) | 2844 (31.4%) | 2715 (31.5%) | 950 (31.8%) | <0.001 |
| Current smoker | 3190 (15.2%) | 1401 (15.3%) | 1294 (14.8%) | 495 (16.3%) | 0.16 |
| Mediterranean Diet Score: 6-9 <sup>◇</sup> | 4069 (26.6%) | 1810 (26.5%) | 1696 (26.7%) | 563 (26.6%) |  |
| Depression: 3-12 <sup>▽</sup> | 3138 (15.1%) | 1317 (14.5%) | 1308 (15.1%) | 513 (17.0%) |  |
| Family MI/stroke history | 12293 (66.1%) | 5237 (63.9%) | 5219 (67.3%) | 1837 (69.2%) | <0.001 |
| PREVENT Base+SDI HF Risk: <3% | 5436 (26.0%) | 3077 (33.6%) | 1968 (22.6%) | 391 (12.9%) | <0.001 |
| 3-<5% | 3656 (17.5%) | 1848 (20.2%) | 1442 (16.5%) | 366 (12.0%) |  |
| 5-<10% | 5759 (27.5%) | 2386 (26.0%) | 2516 (28.8%) | 857 (28.2%) |  |
| >=10% | 6072 (29.0%) | 1852 (20.2%) | 2796 (32.1%) | 1424 (46.9%) |  |

Data are presented as mean (SD) or frequency (percent).
\*Hypertension defined as systolic blood pressure $\geq 130$ mm Hg and/or diastolic blood pressure $\geq 80$ mm Hg or self-reported current use of medication to control blood pressure.
° eGFR was calculated using the Chronic Kidney Disease Epidemiology Collaboration 2021 creatinine equation using standardized or calibrated serum creatinine.
\*Diabetes mellitus defined as fasting glucose $\geq 126$ mg/dL and/or non-fasting glucose $\geq 200$ mg/dL or self-reported current use of medication to control blood sugar.
°Mediterranean Diet Adherence, high score more than 6
▽Depression score calculated using Center for Epidemiological Studies-Depression scale; scores $\geq 3$ indicate risk for clinical depression)
Abbreviations: HDL, high-density lipoprotein, MI, myocardial infarction, PREVENT, Predicting Risk of CVD EVENTS, CVD, cardiovascular disease, eGFR, estimated glomerular filtration rate.

### Risk classification

Table 2 shows the distribution of participants in the 4 PREVENT score categories within each ECG category. Using 3% 10-year risk to define low risk, 26.0% of the sample was classified as low risk, 17.5% as borderline risk (3-<5%), 27.5% as intermediate risk (5%-<10%), and 29.0% as high risk (≥10%). Among those at low/borderline risk i.e., less than 5%, 45.8% had at least one ECG abnormality; 37.5% had only minor ECG abnormality and 8.3% had any major ECG abnormality.

**Table 2:** ECG categories in each PREVENT risk score classification.

|  | All | ECG categories |  |  |  |  |  |
| --- | --- | --- | --- | --- | --- | --- | --- |
|  |  | Normal |  | Minor |  | Major |  |
|  | N (%) | N | row % | N | row % | N | row % |
| **All** | 20923 | 9163 | 43.8 | 8722 | 41.7 | 3038 | 14.5 |
| **PREVENT HF Risk Score** |  |  |  |  |  |  |  |
| **<3%** | 5436 (26.0) | 3077 | 56.6 | 1968 | 36.2 | 391 | 7.2 |
| **3% - <5%** | 3656 (17.5) | 1848 | 50.5 | 1442 | 39.5 | 366 | 10.0 |
| **5% - <10%** | 5759 (27.5) | 2386 | 41.4 | 2516 | 43.7 | 857 | 14.9 |
| **$\geq 10\%$** | 6072 (29.0) | 1852 | 30.5 | 2796 | 46.1 | 1424 | 23.4 |

### Kaplan-Meier survival curves

During follow-up, 6.0% of the sample had a HF event (Figure 1). HF events occurred in 3.3% of the sample with normal ECG, 6.2% of those with only minor abnormalities, and 13.2% of those with any major abnormality. HF events occurred in 0.9% in the <3% low-risk group, 3.1% of the borderline risk group, 6.1% in the intermediate risk group, and 12.1% of the high risk group (Figure 2). Survival curves stratified on sex, race and age are shown in Supplemental Figure 2, 3 and 4, respectively with 4 ECG categories (normal, 1 minor, 2 or more minors and any major abnormalities), showing similar patterns in these groups as in the overall sample.

**Figure 1:**
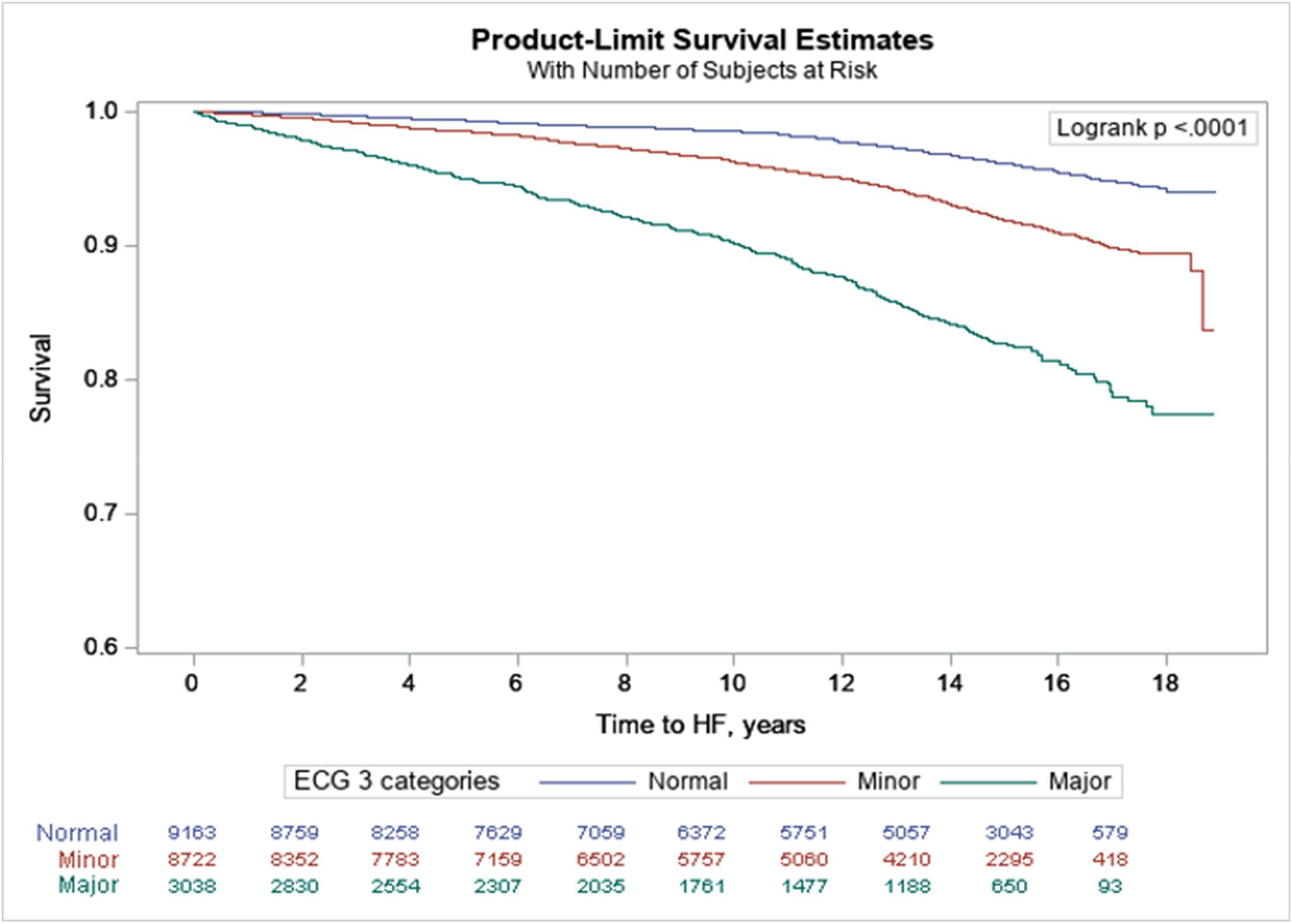
Kaplan-Meier Survival Curves for HF incidents during follow-up (2003-2021) by ECG categories in the overall population.

**Figure 2:**
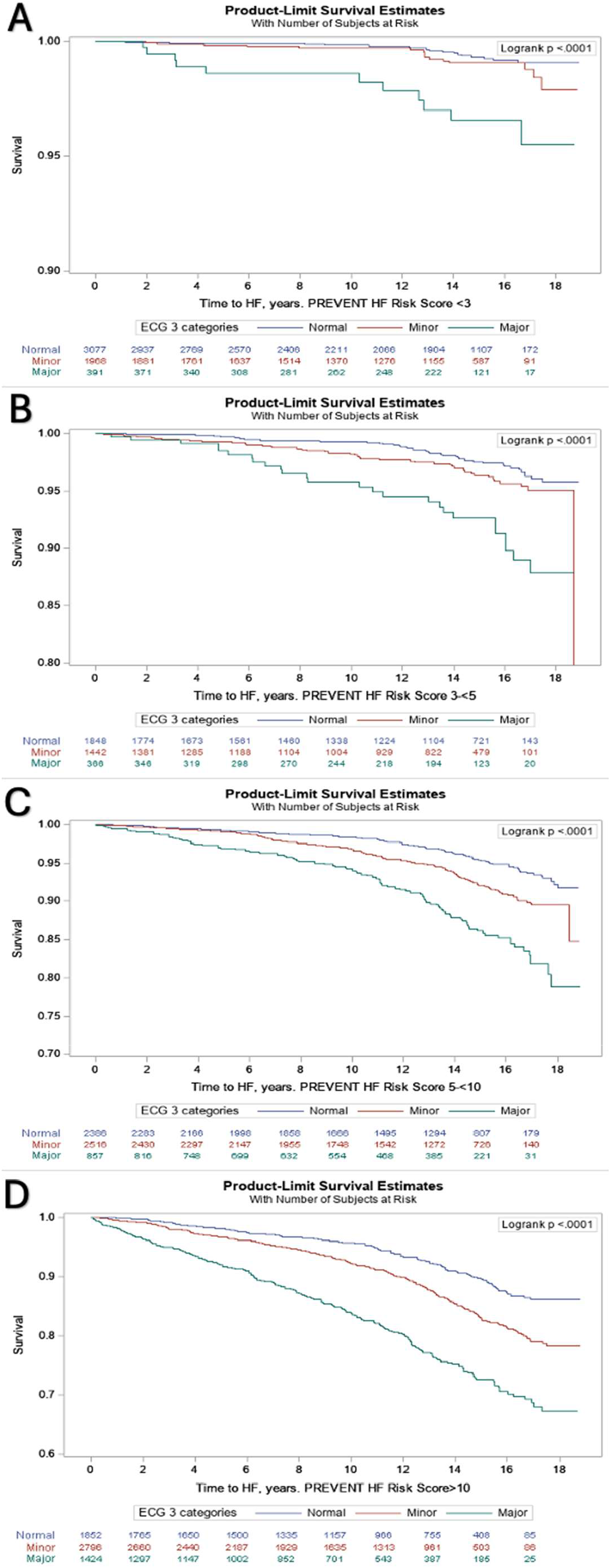
Kaplan-Meier probability of being HF/HF death free during follow-up (2003-2021) by baseline electrocardiogram abnormalities for 3 categories of PREVENT score: A - <3%, B – 3%-<5%, C – 5-<10%, D – ≥10%.

### Multivariable modeling results

The Cox proportional hazard models showed that compared to those with normal ECGs, individuals with any ECG abnormality at baseline had higher risk of HF events in both age-sex adjusted and fully adjusted models (Table 3). The age-sex adjusted model HR for only minor ECG abnormalities was 1.69 (95% CI 1.46-1.95), and for any major ECG abnormality was 3.17 (95% CI 2.71-3.70). The relationship remained after full adjustment; the HR for only minor ECG abnormalities was 1.56 (95% CI 1.35-1.80), and for any major ECG abnormality was 2.56 (95% CI 2.18-3.00).

**Table 3.**
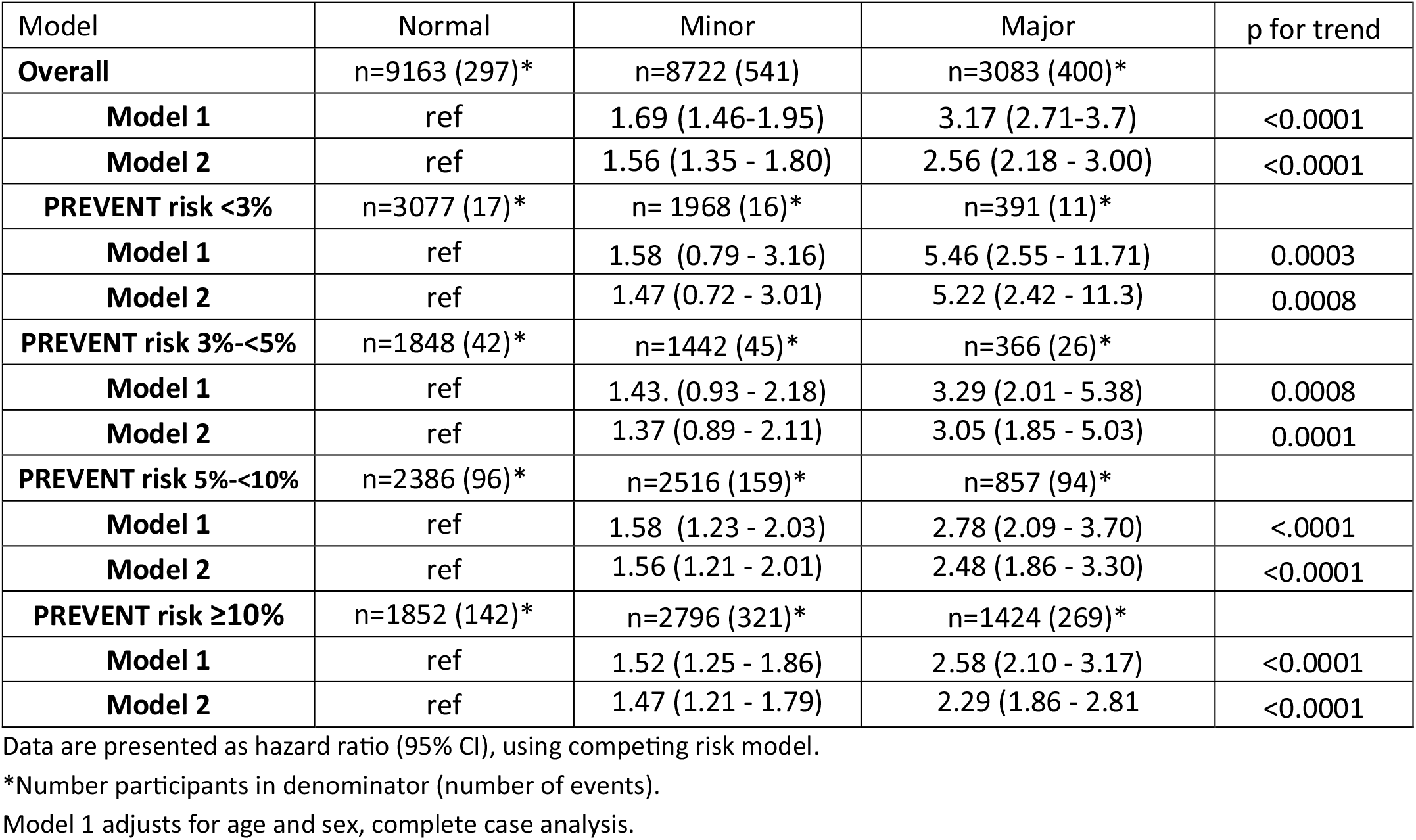
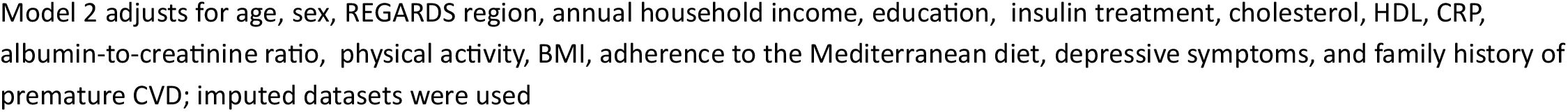
Risk of Incident Heart Failure by ECG Abnormality Categories According to PREVENT HF 10-year risk groups of <3%, 3 – <5%, 5-<10% and ≥10%.

| Model | Normal | Minor | Major | p for trend |
| --- | --- | --- | --- | --- |
| **Overall** | n=9163 (297)* | n=8722 (541) | n=3083 (400)* |  |
| **Model 1** | ref | 1.69 (1.46-1.95) | 3.17 (2.71-3.7) | <0.0001 |
| **Model 2** | ref | 1.56 (1.35 - 1.80) | 2.56 (2.18 - 3.00) | <0.0001 |
| **PREVENT risk <3%** | n=3077 (17)* | n= 1968 (16)* | n=391 (11)* |  |
| **Model 1** | ref | 1.58 (0.79 - 3.16) | 5.46 (2.55 - 11.71) | 0.0003 |
| **Model 2** | ref | 1.47 (0.72 - 3.01) | 5.22 (2.42 - 11.3) | 0.0008 |
| **PREVENT risk 3%-<5%** | n=1848 (42)* | n=1442 (45)* | n=366 (26)* |  |
| **Model 1** | ref | 1.43. (0.93 - 2.18) | 3.29 (2.01 - 5.38) | 0.0008 |
| **Model 2** | ref | 1.37 (0.89 - 2.11) | 3.05 (1.85 - 5.03) | 0.0001 |
| **PREVENT risk 5%-<10%** | n=2386 (96)* | n=2516 (159)* | n=857 (94)* |  |
| **Model 1** | ref | 1.58 (1.23 - 2.03) | 2.78 (2.09 - 3.70) | <.0001 |
| **Model 2** | ref | 1.56 (1.21 - 2.01) | 2.48 (1.86 - 3.30) | <0.0001 |
| **PREVENT risk $\geq 10\%$** | n=1852 (142)* | n=2796 (321)* | n=1424 (269)* |  |
| **Model 1** | ref | 1.52 (1.25 - 1.86) | 2.58 (2.10 - 3.17) | <0.0001 |
| **Model 2** | ref | 1.47 (1.21 - 1.79) | 2.29 (1.86 - 2.81) | <0.0001 |

Data are presented as hazard ratio (95% CI), using competing risk model.
\*Number participants in denominator (number of events).
Model 1 adjusts for age and sex, complete case analysis.
Model 2 adjusts for age, sex, REGARDS region, annual household income, education, insulin treatment, cholesterol, HDL, CRP, albumin-to-creatinine ratio, physical activity, BMI, adherence to the Mediterranean diet, depressive symptoms, and family history of premature CVD; imputed datasets were used

Table 3 also shows HR for models stratified on PREVENT HF 10-year risk by 4 risk categories. We found that individuals classified as having <3% risk had associations with ECG abnormalities as strong or stronger association than those observed in other risk groups: the age- and sex-adjusted HR for only minor ECG abnormalities in the <3% was 1.58 (95% CI 0.79-3.16) and remained strong after full adjustment with HR 1.47 (95% CI 0.72 - 3.01), and the age- and sex-adjusted HR for any major ECG abnormality was 5.46 (95% CI 2.55 - 11.71) and remained strong after full adjustment with HR 5.22 (2.42 - 11.30). In the borderline risk group, the HR for any major ECG abnormality was stronger than the HRs in the intermediate- and high-risk groups, The age- and sex-adjusted model HR was 3.29 (95% CI 2.01 - 5.38) and 3.05 (95% CI 1.85 - 5.03) after full adjustment. See supplementary materials for models with PREVENT HF groups of <7.5%, 7.5%-20, >20%.

The p-value for interactions with age (p=0.53), sex (p=0.17), and race (p=0.12), were not statistically significant suggesting that relationships were similar for race, age, and sex groups.

## DISCUSSION

In this biracial, large, national sample of more than 20,000 participants from the REGARDS study with a median follow-up of more than 13 years, we observed a strong, significant association between baseline ECG abnormalities and incident HF events. Within each group risk of HF based on the PREVENT HF equation, there was a consistent association between each ECG abnormality group and HF events, with greater risk for major compared to minor abnormalities. Interestingly, the association was strongest in the low/borderline risk groups compared to intermediate and high-risk groups, especially for major ECG abnormalities. These results add to growing evidence that ECGs may be useful in further risk stratifying individuals, especially those who would otherwise be classified as low/borderline-risk according to the PREVENT equation.

Preventive strategies are needed to address the growing burden of HF. Although CVD and ASCVD preventive guidelines are well established in the literature,^18^ that is not the case for HF. Prior cardiovascular disease risk assessment tools did not account for heart failure. The landscape of HF prevention started to change with the introduction of the PREVENT equations, which introduced a validated and calibrated tool to assess risk for incident HF.^5^ Accordingly, the 2022 ACC/AHA/HFSA guidelines supported the use of validated multivariable risk scores in the general population to estimate the subsequent risk of HF.^19^ Given the size of this subpopulation of low-/borderline-risk (43.5% of our cohort), it may be important to develop strategies to enhance stratification of this subgroup, as some individuals with this subgroup are likely to be higher risk than others. Baseline ECG abnormalities could serve as such a risk-enhancer. Prior work has shown that the presence of ECG abnormalities are an independent risk factor for HF.^6– 11^ Of note, these studies are outdated and of small sample size. We now extend these findings using explicit ECG criteria among a contemporary cohort to show that this independent risk is present across all PREVENT risk groups. This supports the notion that ECG abnormalities could serve as a risk enhancer and may guide the reclassification of individuals from class A to class B pre-HF.

A practical approach might incorporate the incremental value of ECG could start with a quantitative risk assessment with PREVENT to calculate the 10-year risk of HF events, with further personalization of individual risk based on a baseline ECG. Although the Minnesota Code ECG criteria are complex, there is available software that could theoretically be developed and integrated into ECG machines. Ongoing work shows promise of the ability of large language models (LLMs) in detecting systolic ventricular dysfunction based on ECG abnormalities with deep learning algorithms.^20,21^ LLMs could potentially be trained on large volumes of ECGs to detect the specific ECG abnormalities that increase risk.

Strengths of our study include its large, national, biracial sample, long follow-up period, expert adjudication of events following national guidelines, rigorously collected data, rigorous interpretation of ECGs by a single center, and the high likelihood of generalizability of findings given the national scope. However, the results of our study need to be interpreted cautiously in the context of several limitations. The observational nature of the study warrants caution in drawing causal inferences. Although the REGARDS cohort included a large sample of Black participants, it did not include Latinos and other groups living in the US, warranting studies that do include these populations. As in any research study, participants may not be representative of the general population. Not all participants had an ECG available. Some participants had missing data, which we compensated for by using multiple imputation. Some covariates were self-reported, which may have introduced bias. There is also potential residual confounding due to the observational design, particularly regarding factors such as additional medication use, access to care, etc. These factors may not be fully captured in the models.

In conclusion, we found an association between baseline ECG abnormalities and incident HF events across all PREVENT risk groups. Associations were strongest with major ECG abnormalities, and among those in individuals who would otherwise have been considered low/borderline-risk by the PREVENT equation. Our study supports the potential role of baseline ECG in HF risk stratification.

## Acknowledgment

This research was conducted using data from the REasons for Geographic and Racial Differences in Stroke (REGARDS) study. The authors thank the investigators, staff, and participants of REGARDS for their valuable contributions. A complete list of participating REGARDS investigators and institutions can be found at: https://www.uab.edu/soph/regardsstudy/. The contents do not represent the views of the U.S. Department of Veterans Affairs or the United States Government.

## Statement of Ethics

The REGARDS study was approved by the institutional review boards of all participating institutions. A full list of sites and ethics committees is available at: https://www.uab.edu/soph/regardsstudy/. All participants provided written informed consent.

## Data Availability Statement

The data in this study were obtained from the REGARDS study, a third-party source, where access restrictions apply. Datasets may be requested from the REGARDS Study team at https://www.uab.edu/soph/regardsstudy/

